# Awareness, Attitude, and Perception of Telemedicine Among Trinidad and Tobago Population

**DOI:** 10.1101/2023.12.05.23299482

**Authors:** Ngozika Esther Ezinne, Dipesh Bhattarai, Anayochukwu Edward Anyasodor, Ellen K. Antwi-Adjei, Kingsley Kene Ekemiri, James Andrew Armitage, Uchechukwu Levi Osuagwu

**Affiliations:** Optometry Unit, Department of Clinical Surgical Sciences, University of the West Indies, Trinidad and Tobago; School of Medicine (Optometry), Deakin University, Victoria, Australia; Rural Health Research Institute, Charles Sturt University, Orange, NSW Australia; School of Optometry, University of Alabama at Birmingham, AL, USA; Department of Optometry and Visual Science, Kwame Nkrumah University of Science and Technology, Ghana; School of Medicine, Bathurst Rural Clinical School, Western Sydney University, Bathurst NSW, Australia; African Vision Research Institute, Department of optometry, University of KwaZulu-Natal, Durban South Africa

## Abstract

**Aim:** To assess the awareness, attitude and perception of Trinidad and Tobago population towards telemedicine

**Method:** A cross-sectional study was conducted using a structured questionnaire on patients that visited health centres in the North Central region. A systematic random sampling method was used to select participants. Descriptive statistics including mean and standard deviation were used to calculate categorical and continuous variables. Comparison between the categorical groups of the demographic variables for each of the three main outcome variables were analysed through one-way analysis of variance (ANOVA). Statistical significance was defined as P<0.05.

**Results:** A total of 528 participated in the study. Most (60%) of them were female, and aged 21 to 40 years (62.1%). Awareness of telemedicine was 34.4%, but the majority (82.5%) had never used telemedicine before. About half (51.3%) acknowledged the necessity of telemedicine but few (36.4%) were satisfied with the services. Most (64%) were willing to try mobile-based healthcare apps. Concerns over lack of familiarity with telemedicine platforms (44.5%) and result accuracy (15.5%) were the major barriers to using telemedicine. Awareness of telemedicine was significantly associated with being female (P < 0.001), a medical profession (P = 0.004), familiarity in use of computers (P = 0.004) and frequent interaction with doctors online (P < 0.001). Positive attitude towards telemedicine was associated with having a diploma, being a medical professional, being computer literate and frequent interaction with doctor online. Positive perception towards telemedicine was associated with marital status (Single or Previously Married) (P = 0.011), one’s ability to use the computer (P = 0.009), their level of competency in computer usage (P = 0.002), and frequency of interacting with doctors online (P < 0.002).

**Conclusion:** The study revealed that although the level of telemedicine awareness is low, the majority of respondents demonstrated positive attitude and perception towards telemedicine. The findings suggest the need to educate the public on the benefits of telemedicine and create awareness of its use in T&T.

## Introduction

Telemedicine, a change in paradigm in healthcare, leverages technology including text messages, emails, telephone or audio and video consultation to provide remote health care services [1]. Telemedicine refers to the usage of technology such as internet, wireless technology, satellite and telephone media to deliver health care remotely [2]. Depending on the available telecommunication technology and the infrastructure, telemedicine occurs synchronously in real-time between a patient and a provider or asynchronously where images, test results and information are captured, stored and forwarded from one location to another for later review by the provider [3,4].

Prior to shifts induced by the COVID-19 pandemic, telemedicine, although promising, remained underutilized in many regions [5]. The primary role of telemedicine was mainly to offer remote consultations, especially for underserved rural populations [6]. Across specialties such as optometry, mental health, paediatrics, and more, telemedicine’s offers a new era of healthcare delivery [2,7–9]. Notably, the advantages of telemedicine go beyond convenience to transformative solution for the healthcare sector that are potentially cost-effective and accessible during an epidemic [8,10–13].

The emergence of the COVID-19 pandemic reshaped global healthcare landscapes and accelerated the adoption of telemedicine as a necessity rather than a choice [14,15]. In the face of global uncertainties, telemedicine became a crucial tool for continuity by providing a lifeline to medical care while minimizing viral transmission risks [16]. In addition, telemedicine offered a solution to the challenges posed by restrictions on physical consultations, limited healthcare infrastructure, and a heightened need for minimizing human interactions [6]. While telemedicine emerged as a valuable tool, it also brought several core challenges including technological issues with data security and privacy [17].

Amidst these advancements, a distinct disparity emerged. While telemedicine found its foothold across continents, the role of telemedicine in the Caribbean remains undocumented [18]. Unveiling the potential drivers and barriers for its acceptance in specific regions, such as Trinidad and Tobago (T&T), has now gained paramount importance in the post-COVID-19 era. T&T, with its unique sociocultural dynamics drawn from its multiracial population and free health care system, makes the country an interesting place to explore the adoption, awareness, and perception of telemedicine. Furthermore, the fact that its neighbouring countries have made significant progress in telemedicine practices including formulation of guidelines and policies [1], makes it important to study the situation in T & T. A study of the utility of telemedicine among Jamaicans [19] showed that the majority preferred face-to-face consultation to telemedicine which may be due to lack of awareness about the benefits of telemedicine [1]. Evaluating telemedicine awareness and perception is key to ensuring that current healthcare strategies align with evolving patient preferences and technological advancements.

Telemedicine helps to navigate the challenges by providing alternatives for patients who may not want an in-person consultation with a healthcare provider to receive care [13,20,21]. Its use is essential at bridging the gap in health care delivery at times when face to face consultation may not be possible such as during a disease outbreak, an epidemic, or a pandemic. As the pandemic continues to impact healthcare delivery, the readiness of societies to embrace telemedicine as a viable alternative to traditional healthcare becomes an imperative question.

This study seeks to bridge the knowledge gap and provide insights into the awareness and perception of telemedicine in T&T. By understanding the factors that shape the reception of telemedicine among the public, this research aims to provide evidence for healthcare policymakers, providers, and technologists to integrate telemedicine more seamlessly into the evolving healthcare landscape. Past studies [9,18] awareness, knowledge, and perception of telemedicine have focused on health care practitioners with little or no evidence of what the public the telemedicine users thinks about it. Moreso, in T & T, there is a paucity of information on telemedicine both from the practitioners’ perspective and from the public. This study was therefore designed to provide evidence from the public perspective about their level of awareness, knowledge and how their perception of telemedicine practice in T&T. Findings from this study will provide an understanding of what the public thinks about telemedicine practice in T&T, provide evidence that can be used for comparison with other studies, and for policy decision making by the relevant government agencies when designing programs around telemedicine.

## Materials and methods

### Study design and setting

This cross-sectional study was conducted in Trinidad and Tobago, a twin island nation in the Southeastern West Indies, with a population of 1,363,985. The country has 101 health centres that offer free primary healthcare services, administered by five regional health authorities. This study focused on the North Central Regional Health Authority (NCRHA), which operates 15 health centres. The method of selection of this authority to ensure its representativeness is described below.

### Study Population

Participants included all individuals aged 18 years and above who sought healthcare services at NCRHA health centres. Participants were recruited between 10^th^ August 2021 and 30^th^ December 2021. Inclusion criteria were adults aged 18 years and older, who visited selected health centres and provided voluntary consent. Those who were critically ill during data collection were excluded.

### Sample size

The sample size was determined using a single-proportion population formula. With a confidence level of 95% (*Z_α/2_* = 1.96), an estimated awareness and perception proportion of 50%, and a 5% margin of error, the sample size was calculated as 385. Considering non-response rates, the final sample size was doubled to 770.

### Sampling technique

Five health centres were randomly chosen from the 15 in NCRHA. The total sample size was divided by 5 to allocate participants to each centre. Systematic random selection was employed, choosing every third patient from the register. In case of unavailability or unwillingness to participate in the study, the next patient on the list was selected.

### Data collection

A validated questionnaire used in similar studies in India and the Philippines [22,23] was adapted after modification to contextualise for the T&T population and face validity was verified by pretesting on 20 persons not part of the study. The final questionnaire which is presented as a supplementary file (S1 Appendix) covered items including: demography (age groups, sex, marital status, religion, ethnicity, residence, educational status, occupation and income which was grouped according to salary/pay scales in T & T, computer use variable (ability to use computer, level of proficiency in use of computer; type of device used), presence of chronic disease, and frequency of interaction with doctor. Two trained optometry students acted as research assistants, collecting face-to-face data during clinic days and assisted the participants to complete the questionnaire where necessary. Supervision and double data entry were employed to ensure accuracy, and data cleaning was performed prior to analysis. The number of completed questionnaires from each centre is shown in Supplementary file 2 (S2 Appendix).

### Ethical Considerations

The study adhered to the Declaration of Helsinki. Approval was obtained from the Research and Ethics Committee of the University of the West Indies, Saint Augustine campus (CREC-SA/0633/11/2020). Permissions were also secured from health centre administrators. Participants were provided with information and reasons to participate, and written informed consent was obtained. Confidentiality was maintained, and COVID-19 protocols were followed.

### Data analysis

Descriptive statistics were employed, presenting proportions for categorical variables and means (with standard deviations, SD) for continuous variables. In the questionnaire, ‘Yes’ responses were scored as 1 and ‘No’ responses as 0. For perception items, scores ranged from 0 (’strongly disagree’) to 4 (’strongly agree’). ‘Neutral’ responses were assigned a score of 2 for perception items. Raw scores for each item were converted to percentages, checked for normality, and correlated using the Pearson correlation coefficient. The calculated mean percentage scores were compared using paired t-tests between the three domains. Comparison between the categorical groups of the demographic variables for each of the three main outcome variables (awareness, attitude, and perception) were analysed through one-way analysis of variance (ANOVA). Statistical significance was set at P<0.05. Further analysis was conducted to explore this relationship between participants perceived satisfaction and residence. All data collected were exported to Statistical Package for Social Sciences for Windows, version 29 (IBM Corp., Armonk, N.Y., USA).

## Results

A total of 528 participants successfully completed the survey, mostly females (60.0%) and 62.1% were aged between 21 and 40 years. The majority (60.4%) were not married at the time of this study. Among the participants, 77.4% identified as Christians, 54.5% were of Afro-Trinidad ethnicity, and 51.5% resided in the peri-urban areas of Trinidad and Tobago. Regarding educational attainment, 46.5% held a diploma as their highest qualification, and 92.7% were employed in non-medical services. Regarding self-reported health condition, 82.1% indicated the absence of chronic diseases, whereas 17.9% reported having chronic conditions. Proficiency in computer usage was affirmed by 91.8% of participants, with 58.1% self-rating their skills as intermediate. In terms of online medical interactions, only 10.1% reported frequent engagement with doctors, while 46.4% reported never having interacted with doctors online. Further details of the characteristics are shown in Table 2.

**Table 2:** Socio-demographic characteristics of the study participants (N=528).

| Variable | Subgroup | Frequency (%) |
| --- | --- | --- |
| Gender | Female | 315 (59.7) |
|  | Male | 210 (39.8) |
| Age group, years | Less than 20 | 18 (3.4) |
|  | 21-40 | 328 (62.1) |
|  | 41-60 | 128 (24.2) |
|  | Above 61 | 54 (10.2) |
| Marital status | Single or Previously Married | 319 (60.4) |
|  | Married | 209 (39.6) |
| Religion | Christian | 408 (77.4) |
|  | Non-Christian | 119 (22.6) |
| Ethnicity | Afro-Trinidad | 287 (54.5) |
|  | Indo-Trinidad | 68 (12.9) |
|  | Mixed | 164 (31.1) |
|  | Others | 8 (1.5) |
| Residence | Urban | 167 (32.0) |
|  | Peri-urban | 269 (51.5) |
|  | Rural | 86 (16.5) |
| Highest educational qualification | Diploma or below | 239 (46.5) |
|  | Bachelors | 201 (39.1) |
|  | Post-graduate | 74 (14.4) |
| Occupation | Non-medical | 420 (92.7) |
|  | Medical | 33 (7.3) |
| Employment status | Public | 137 (26.6) |
|  | Non-public | 277 (53.7) |
|  | Unemployed | 102 (19.8) |
| Income status<sup>†</sup> | Poor <3000.00 TTD (Minimum wage) | 18 (3.4) |
|  | Not so good (3000 to 5000 TTD) | 50 (9.5) |
|  | Average/reasonable (6000 to 10, 000 TTD) | 319 (60.9) |
|  | Good (11,000 to 29, 000 TTD) | 129 (24.6) |
|  | Very good (30, 000 and above) | 8 (1.5) |
| Presence of chronic disease | Yes | 94 (17.9) |
|  | No | 431 (82.1) |
| Ability to use computer effectively | Yes | 484 (91.8) |
|  | No | 43 (8.2) |
| Level of use of computer | Beginner | 27 (5.1) |
|  | Intermediate | 306 (58.1) |
|  | Professional | 194 (36.8) |
| Smart device used. | Phones | 266 (54.2) |
|  | Laptops/Tablets | 41 (8.4) |
|  | Both | 184 (37.5) |
| Frequency of interacting with your doctor online | Frequent (At least once in a week) | 52 (10.1) |
|  | Seldom (Less than once in a month) | 225 (43.5) |
|  | Never | 240 (46.4) |

<sup>†</sup>= Grouped according to salary or pay scales in Trinidad and Tobago

The participants’ perceptions of telemedicine are shown in Table 3. Approximately half (51.3%) acknowledged the necessity of telemedicine, while 42.5% were neutral. Only about one-third (36.4%) expressed satisfaction with telemedicine services, and 47.3% preferred in-person hospital visits during a pandemic. Notably, around two-thirds (64%) expressed willingness to use telemedicine in the future. Moreover, 60.8% agreed that telemedicine is a viable pathway for medical care, and 69.8% agreed that telemedicine can save time and money. Responses of participants on whether they were satisfied with the use of telemedicine revealed that 36.2% were satisfied, 17.3% dissatisfied and 46.5% remained neutral. Their responses differed significantly with residence status such that significantly more people in the urban (38.9%) and peri-urban regions (37.4%) expressed satisfaction with the use of telemedicine compared to fewer people (28.2%; P=0.013) in the rural regions.

**Table 3:** Participant responses on awareness, attitude and perception towards telemedicine in Trinidad and Tobago (N=528).

| Variables | Frequency (%) |
| --- | --- |
| **Awareness of telemedicine** |  |
| Yes | 180 (34.3) |
| No | 343 (65.6) |
| **Use of telemedicine** |  |
| Yes | 91 (17.5) |
| No | 430 (82.5) |
| **Aware of any telemedicine platform** |  |
| Yes | 48 (9.2) |
| No | 472 (90.8) |
| **Correctly named at least one telemedicine platform** |  |
| Yes | 45 (10.2) |
| No | 474 (89.8) |
| **Attitude** |  |
| **Are you concerned with privacy issues or confidentiality of your information if you interact with your doctor online?** |  |
| Yes | 174 (33.8) |
| No | 233 (45.2) |
| Maybe | 108 (21.0) |
| **Would you be willing to try mobile based health care app?** |  |
| Yes | 335 (64.4) |
| No | 26 (5.0) |
| Maybe | 159 (30.6) |
| **Please indicate mode of telemedicine you would prefer.** |  |
| Video consultation | 156 (30.5) |
| Audio consultation | 21 (4.1) |
| Text based consultation | 10 (2.0) |
| Combination of any of the two | 210 (41.0) |
| All the above | 115 (22.5) |
| **Perception** |  |
| **Do you agree on the necessity of telemedicine?** |  |
| Strongly disagree | 16 (3.1) |
| Disagree | 15 (3.0) |
| Neutral | 216 (42.5) |
| Agree | 148 (29.1) |
| Strongly agree | 113 (22.2) |
| **Satisfied with telemedicine** |  |
| Strongly disagree | 21 (11.2) |
| Disagree | 11 (5.9) |
| Neutral | 87 (46.5) |
| Agree | 40 (21.4) |
| Strongly agree | 28 (15.0) |
| **Better preference to going to hospital during a pandemic** |  |
| Strongly disagree | 62 (12.8) |
| Disagree | 54 (11.1) |
| Neutral | 140 (28.8) |
| Agree | 129 (26.5) |
| Strongly agree | 101 (20.8) |
| **Willing to use telemedicine in the future** |  |
| Strongly disagree | 11 (2.2) |
| Disagree | 15 (3.0) |
| Neutral | 149 (29.7) |
| Agree | 172 (34.3) |
| Strongly agree | 154 (30.7) |
| **Telemedicine is a viable approach for providing medical care services to patients** |  |
| Strongly disagree | 19 (3.7) |
| Disagree | 11 (2.2) |
| Neutral | 170 (33.3) |
| Agree | 242 (47.5) |
| Strongly agree | 68 (13.3) |
| **Use of telemedicine system can save time and money** |  |
| Strongly disagree | 20 (3.9) |
| Disagree | 11 (2.2) |
| Neutral | 123 (24.1) |
| Agree | 272 (53.2) |
| Strongly agree | 85 (16.6) |

Table 3 also outlines participants’ attitudes towards telemedicine and their preferred mode of use. Table 4 illustrates the perceived barriers to using telemedicine. A reasonable proportion (33.8%) expressed concerns regarding online privacy and confidentiality when sharing personal information. Conversely, 45.2% were not concerned about this issue. The majority (64.4%) indicated willingness to try mobile-based healthcare apps. Concerns over lack of familiarity with the technique (44.5%) and result accuracy (15.5%) emerged as the primary barriers to using telemedicine.

**Table 4:** Perceived barriers among participants in using telemedicine.

| Barriers to using telemedicine | Frequency (%) |
| --- | --- |
| Unfamiliar with the technique | 230 (44.5) |
| Concern with accuracy of the result | 80 (15.5) |
| Privacy and security concern | 74 (14.3) |
| Not interested | 42 (8.1) |
| Cost | 28 (5.4) |
| Policy barrier | 22 (4.3) |
| Unavailability of trained staff | 12 (2.3) |
| No guideline for use | 8 (1.5) |
| Poor internet | 7 (1.4) |
| Others | 14 (2.7) |

Table 5 shows the results of one-way ANOVA comparing the mean attitude and perception scores towards telemedicine for participants by their demographic variables. Significant differences in mean awareness scores towards telemedicine were observed between genders, with females having higher scores (1.60 [95%CI: 1.46, 1.73]) than males (1.25 [95%CI:1.13, 1.37], P < 0.001). Similarly, medical professionals showed greater attitude scores towards telemedicine compared to non-medical professionals (2.03 [95%CI:1.55, 2.51] vs 1.45 [95%CI:1.35, 1.55], P=0.004). Those proficient in the use of computer (1.50 [95%CI:1.40, 1.60] vs 1.00 [95%CI:0.70, 1.30], P=0.004) and advanced computer users (1.65 [95%CI:1.50, 1.81] vs 1.07 [95%CI:0.66, 1.48], P=0.004) exhibited higher attitude scores. Participants frequently interacting with doctors online (2.40 [95%CI:2.04, 2.76]) scored higher on attitude compared to those interacting seldom (1.61 [95%CI:1.46, 1.76]) or never (1.16 [95%CI:1.05, 1.27], P<0.001).

**Table 5:** Mean scores [95% confidence intervals, CI] for awareness and perception of telemedicine among Trinidad and Tobago population. (N=385).

| Subgroups | Awareness<br>Mean [95% CI] | P-value | Perception<br>Mean [95% CI] | P-value |
| --- | --- | --- | --- | --- |
| **Gender** |  |  |  |  |
| Female | 1.60 [1.46, 1.73] | **<0.001** | 3.52 [3.45, 3.59] | 0.608 |
| Male | 1.25 [1.13, 1.37] |  | 3.49 [3.42, 3.57] |  |
| **Age group in years** |  |  |  |  |
| Less than 20 | 1.28 [0.80, 1.75] | 0.691 | 3.65 [3.35, 3.96] | 0.075 |
| 21-40 | 1.47 [1.35, 1.60] |  | 3.55 [3.49, 3.61] |  |
| 41-60 | 1.51 [1.31, 1.70] |  | 3.40 [3.28, 3.52] |  |
| Above 61 | 1.33 [1.08, 1.59] |  | 3.48 [3.33, 3.63] |  |
| **Marital status** |  |  |  |  |
| Single or Previously Married | 1.48 [1.35, 1.60] | 0.678 | 3.56 [3.50, 3.63] | **0.011** |
| Married | 1.44 [1.29, 1.58] |  | 3.42 [3.34, 3.51] |  |
| **Religion** |  |  |  |  |
| Christian | 1.52 [1.41, 1.63] | 0.021 | 3.52 [3.45, 3.57] | 0.736 |
| Non-Christian | 1.25 [1.06, 1.44] |  | 3.49 [3.38, 3.61] |  |
| **Ethnicity** |  |  |  |  |
| Afro-Trinidad | 1.38 [1.26, 1.50] | 0.277 | 3.51 [3.44, 3.58] | 0.760 |
| Indo-Trinidad | 1.62 [1.31, 1.93] |  | 3.58 [3.43, 3.72] |  |
| Mixed | 1.52 [1.35, 1.69] |  | 3.50 [3.40, 3.59] |  |
| Others | 1.75 [0.59, 2.91] |  | 3.40 [2.84, 3.97] |  |
| **Residence** |  |  |  |  |
| Urban | 1.49 [1.31, 1.66] | 0.975 | 3.54 [3.45, 3.63] | 0.579 |
| Peri-urban | 1.46 [1.33, 1.59] |  | 3.48 [3.41, 3.55] |  |
| Rural | 1.48 [1.24, 1.71] |  | 3.52 [3.38, 3.67] |  |
| **Highest educational qualification** |  |  |  |  |
| Diploma or below | 1.35 [1.22, 1.48] | **0.001** | 3.46 [3.38, 3.54] | 0.062 |
| Bachelors | 1.47 [1.31, 1.63] |  | 3.52 [3.45, 3.59] |  |
| Post-graduate | 1.89 [1.62, 2.16] |  | 3.65 [3.49, 3.80] |  |
| **Occupation** |  |  |  |  |
| Medical | 2.03 [1.55, 2.51] | **0.004** | 3.53 [3.47, 3.59] | 0.445 |
| Non-medical | 1.45 [1.35, 1.55] |  | 3.61 [3.41, 3.82] |  |
| **Employment status** |  |  |  |  |
| Public | 1.45 [1.25, 1.65] | 0.935 | 3.54 [3.43, 3.64] | 0.701 |
| Private | 1.48 [1.35, 1.61] |  | 3.50 [3.43, 3.57] |  |
| Unemployed | 1.43 [1.23, 1.63] |  | 3.47 [3.36, 3.59] |  |
| **Income status** |  |  |  |  |
| Poor <3000.00 TTD (Minimum wage) | 1.36 [0.90, 1.85] | 0.287 | 3.31 [2.81, 3.82] | 0.071 |
| Not so good (3000 to 5000 TTD) | 1.10 [0.87, 1.33] |  | 3.43 [3.30, 3.55] |  |
| Average/reasonable (6000 to 10, 000 TTD) | 1.29 [1.18, 1.39] |  | 3.52 [3.45, 3.58] |  |
| Good (11,000 to 29, 000 TTD) | 1.42 [1.27, 1.57] |  | 3.51 [3.41, 3.62] |  |
| Excellent (30, 000 and above) | 1.50 [0.93, 2.07] |  | 4.02 [3.58, 4.46] |  |
| **Presence of chronic disease** |  |  |  |  |
| Yes | 1.66 [1.39, 1.93] | 0.053 | 3.49 [3.35, 3.63] | 0.700 |
| No | 1.42 [1.32, 1.52] |  | 3.51 [3.46, 3.57] |  |
| **Ability to use computer effectively** |  |  |  |  |
| Yes | 1.50 [1.40, 1.60] | **0.004** | 3.53 [3.48, 3.58] | **0.009** |
| No | 1.00 [0.70, 1.30] |  | 3.28 [3.04, 3.51] |  |
| **Level of use of computer** |  |  |  |  |
| Beginner | 1.07 [0.66, 1.48] | **0.003** | 3.17 [2.90, 3.44] | **0.002** |
| Intermediate | 1.37 [1.24, 1.50] |  | 3.49 [3.42, 3.55] |  |
| Professional | 1.65 [1.50, 1.81] |  | 3.59 [3.50, 3.67] |  |
| **Smart device used** |  |  |  |  |
| Phones | 1.37 [1.24, 1.49] | **0.013** | 3.54 [3.47, 3.61] | 0.788 |
| Laptops/Tablets | 1.44 [1.10, 1.78] |  | 3.47 [3.27, 3.67] |  |
| Both | 1.68 [1.51, 1.85] |  | 3.53 [3.44, 3.62] |  |
| **Frequency of interacting with your doctor online** |  |  |  |  |
| Frequent | 2.40 [2.04, 2.76] | **<0.001** | 3.76 [3.62, 3.91] | **<0.001** |
| Seldom | 1.61 [1.46, 1.76] |  | 3.54 [3.46, 3.61] |  |
| Never | 1.16 [1.05, 1.27] |  | 3.43 [3.35, 3.50] |  |

Bolded *P* values represent statistically significant subgroups using one-way analysis of variance.

Perception scores towards telemedicine also exhibited significant variation based on marital status (Single or Previously Married 3.56 [95%CI: 3.50, 3.63] vs Married: 3.42 [95%CI: 3.34, 3.51]; P = 0.011), computer usage proficiency (Yes 3.53 [95%CI: 3.48, 3.58] vs No 3.28 [95%CI: 3.04, 3.51]; P = 0.009), computer usage level (Professional 3.59 [95%CI: 3.50, 3.67] vs Intermediate 3.49 [95%CI: 3.42, 3.55] vs Beginner 3.17 [95%CI: 2.90, 3.44] P = 0.002), and frequency of interacting with doctors online (Frequent 3.76 [95%CI: 3.62, 3.91] vs Intermediate 3.54 [95%CI: 3.46, 3.61] vs Beginner 3.43 [95%CI: 3.35, 3.50] P < 0.002).

## Discussion

Telemedicine has been in existence for over a decade, but the awareness and utilization of telemedicine has not been well covered especially in developing countries including Trinidad and Tobago. COVID-19 pandemic led to a surge in the use of telemedicine in developed countries as an alternative for individuals seeking health care services. Awareness, attitude, and perception of telemedicine have not been previously assessed in a Caribbean population. To the best of our knowledge, our study was the first to assess the awareness and perception of telemedicine among Trinidad and Tobago population. There was an overall poor awareness of telemedicine, its platforms and its utilization. Most of the participants showed a positive attitude, were willing to try a mobile based app or other forms of telemedicine and were not so concerned about the confidentiality of the information obtained. On the other hand, many see it as a necessity, and a viable approach to medical services and are willing to try it in the future. However, a good proportion are not satisfied with it and do not see it as a better choice than going to a hospital. Awareness of telemedicine was significantly associated with being female, a medical professional, and being a user of smartphones. Also, awareness, positive attitude and perception towards telemedicine were significantly associated with the ability to use computer, level of computer skill and frequency of online interaction. Unfamiliarity with the telemedicine techniques and concern with accuracy of results obtained were the major barriers to adoption of the telemedicine by the public. Other findings from the study are discussed below.

### Awareness of telemedicine

The level of awareness of telemedicine recorded in our study is lower than findings from a previous study [9] among optometrists in T&T. It is also lower than 57.4%, 58.7% and 70.1% recorded in Ethiopia [24], Nigeria [25] and Pakistan [8] respectively. The poor awareness of telemedicine recorded in our study could be because telemedicine is still at its early stage in T&T, hence not popular among health care service providers. This is a cause for concern, especially given the substantial surge in new technologies during the recent COVID-19 pandemic [26,27]. This underscores the imperative for government agencies to raise awareness concerning the significance of telemedicine in disease management for a successful adoption and utilization by the population [28]. Other factors are absence of regulatory frameworks governing telemedicine, compounded by limited integration of telemedicine into the healthcare system, and lack of policies addressing the digital gap.

Another notable finding is the infrequent use of telemedicine recorded in the present study which is consistent with findings from other developing countries [9,29,30], despite reports of surge in the use of telemedicine recorded in some developed countries [31,32]. This trend aligns with the poor awareness observed in our study and could be linked to ineffective adoption strategies. It is not surprising that awareness of telemedicine was significantly associated with females in the present study as studies [33,34] revealed that females are more digitally literate because they use phones more than males. On the other hand, those in medical profession are expected to be more aware of telemedicine as they are involved with different forms of medical care, hence will tend to get more information or alternatives to getting best care for their patients thereby exposing them to telemedicine than others.

### Attitude

Our findings showed that T&T population have a favourable attitude towards telemedicine as the majority are willing to try mobile based health app and are not concerned about confidentiality issues raised as a major barrier in the utilization in previous studies [35,36]. Similar findings were recorded in studies on Jordanian [30] and Chinese populations [32]. A positive attitude is essential for a new technology to be adopted and used by the population, therefore there is a need to address this barrier.

### Perception

Although, majority of our study participants agreed that telemedicine is a necessity and a viable health care service that can save time, they are not satisfied with the service and do not perceive it to be a better option than going to a hospital contrary to other study findings [37–40]. This could be because majority of the respondents (51.5%) were residence in peri-urban areas, where there is transportation and easy access to a hospital. Similarly a study had reported that telemedicine is not convenient for patients with better access to care [4]. A study in rural Alabama where there is lack of transportation and specialist care reported higher satisfaction among the participants after telemedicine encounter [3,4], probably because healthcare in the US is not necessarily free compared with T&T.

Negative attitudes towards telemedicine could impede telemedicine adoption, diminishing its potential to enhance healthcare accessibility and efficiency. Furthermore, training on various telemedicine modes and techniques are highly recommended to enhance understanding of telemedicine and its associated health benefits [37,41–43] which will necessitate the development of strategies and investment in continuous professional development for healthcare providers in virtual care [44]. In addition, the fact that positive perception of telemedicine is associated with the use of computer and level of computer skill is expected as those are the basic things essential for an adoption or utilization of telemedicine and are recorded in previous studies [30,35] as reasons for poor awareness and reception of telemedicine. Consequently, policies should prioritize increasing telemedicine perception to encourage the public to maximize its benefits especially in natural disasters or emergencies.

People were generally satisfied (albeit many were neutral) with the use of telemedicine which was consistent with previous studies [45–47]. However, significantly more people in the urban and peri urban regions expressed satisfaction with the use of telemedicine compared with rural residents. This finding was consistent with previous study [47] which reported that rural residence in addition to factors such as educational attainment, prior Internet use were the main predictors that increase the likelihood of being amenable to telemedicine. This higher satisfaction with telemedicine use among urban residents may be related to factors such as better accessibility to technology and infrastructure, where Urban regions typically have better access to high-speed internet, reliable telecommunications networks, and advanced technology infrastructure, which are crucial for the smooth functioning of telemedicine services. On the other hand, rural areas often face challenges related to poor internet connectivity and technological resources, making it difficult for them to fully utilize telemedicine services.

### Barriers

Despite the global proliferation of telemedicine, numerous challenges persist [2]. These highlighted barriers such as technological unfamiliarity, concern for privacy and accuracy of results recorded in the current study. It is evident that performance, encompassing result accuracy hinders telemedicine adoption [48]. However, it should be noted that privacy risks do not consistently deter the adoption of technology-driven healthcare [22]. Other recorded barriers in previous studies were resistance to change, high cost of technology, underdeveloped infrastructure, shortage of technical staff expertise, healthcare provider resistance to change, patient’s resistance to change, lack of training on information technology, cultural aspects, legal issues, patient’s age, and patient’s education level [35,48].

Telemedicine has merits and demerits but the advantages outweigh the disadvantages. There is a potential market for telemedicine in Trinidad and Tobago if more awareness of the service is created. There is therefore a need to establish policies and strategies for easy adoption and utilization of telemedicine in Trinidad and Tobago as adoption of any new technology depends on understanding of its concept by users, obtaining its required skills and the suitable working environment. Moreover, establishing platforms and organizing telemedicine workshops as well as use of secured high-quality audio and video are necessary for tackling barriers with the usage.

### Strengths and limitations

The strength of our study is the fact that our study added new and important information to the literature and is the first to assess the awareness, attitude, and perception of telemedicine in a Caribbean population. Also, the study assesses these participants’ predisposition for telemedicine prior to implementation. Additionally, the study had an adequate sample size sufficient to give an insight about telemedicine in T&T. Nevertheless, our study is not without limitations that should be acknowledged. Firstly, the study employed a cross-sectional study design, which precludes the investigation of causal relationships. Also, the use of questionnaire and close-ended questions could have caused certain participants’ responses to be missed as study participants are subjected to bias.

## Conclusion

The study revealed that although the level of telemedicine awareness is low, majority demonstrated positive attitude and perception towards telemedicine. Therefore, there is a need for more telemedicine awareness and education campaign for the T&T population especially those in the rural areas.

## Data Availability

All relevant data are within the manuscript and its Supporting Information files.

## Acknowledgement

We thank the University of the West Indies, Saint Augustine campus academic research grant for their support during data collection.

## Supporting information

S1 Appendix: Sample of questionnaire used in the study

S2 Appendix: Questionnaire distribution

S2 Appendix: Raw data for telemedicine study in Trinidad and Tobago

